# Features of an aseasonal 2021 RSV epidemic in the UK and Ireland: analysis of the first 10,000 patients

**DOI:** 10.1101/2022.03.03.22271525

**Authors:** D Roland, TC Williams, MD Lyttle, R Marlow, PC Hardelid, I Sinha, OV Swann, A Maxwell-Hodkinson, S Cunningham, the REspiratory Syncytial virus Consortium in EUrope (RESCEU) investigators and the Paediatric Emergency Research United Kingdom and Ireland (PERUKI) Network

**Author notes:** **Corresponding Author:** Damian Roland, SAPPHIRE Group, Health Sciences, Leicester University, Leicester, UK, Paediatric Emergency Medicine Leicester Academic (PEMLA) Group, Children’s Emergency Department, Leicester Royal Infirmary, Leicester, UK.

## Abstract

BronchStart is a prospective cohort study of infants with clinical bronchiolitis attending Emergency Departments in the United Kingdom and Ireland. We found the 2021 summer lower respiratory tract infection peak, although temporally disrupted and with an attenuated disease burden, predominantly affected younger age groups as in previous years.

## Main text

Respiratory syncytial virus (RSV) causes annual winter epidemics that usually peak in December in the UK and Ireland. Infants are disproportionately affected, with infection leading to lower respiratory tract disease, most commonly bronchiolitis, that often result in emergency department visits and hospitalisations. Non-pharmaceutical interventions (NPIs) introduced globally to limit the spread of severe acute respiratory syndrome coronavirus 2 (SARS-CoV-2) led to disruption of the typical RSV seasonality. Australia experienced a delayed RSV peak [1] and a shift in age distribution to older children, with New Zealand reporting more hospital admissions than in previous years [2]. Proposed reasons include larger populations of RSV-naïve infants/children, and decay in transferred maternal RSV IgG antibody. Studies examining the aseasonal resurgence of RSV have been limited by sample size, and lack of information on secondary care episodes and clinical features. The BronchStart study, which commenced in June 2021, is a prospective multi-centre cohort study. Paediatric emergency departments (PED) within PERUKI (Paediatric Emergency Research in the UK and Ireland) submit data on all children under 2 years of age who visit a PED with symptoms of an acute lower respiratory tract infection (diagnosed as bronchiolitis, lower respiratory tract infection, or first episode of acute wheeze), to a secure online Research Electronic Data Capture (REDCap) database. Follow-up information is submitted 7 days later, and study data is made available on a live online dashboard hosted by Microreact [3].

We present initial data for 10,347 infants and children from 44 study sites for the period 1^st^ June to 5^th^ December 2021. The aseasonal 2021 RSV epidemic in the UK has now come to an end, with infections having peaked in August (Figure 1A, [4]). Comparing the age distribution of hospitalised infants <12 months to previous years at two large paediatric centres currently participating in the BronchStart Study (Leicester Children’s Hospital and Bristol Royal Hospital for Children), we observed a similar age distribution (Figure 1B). This suggests reduced community exposure to RSV during the 15 months preceding the start of the season did not result in a clinically significant lack of protective maternal antibody transfer to those <3 months of age, or that the NPIs introduced were not strong enough to prevent low level transmission.

**Figure 1:**
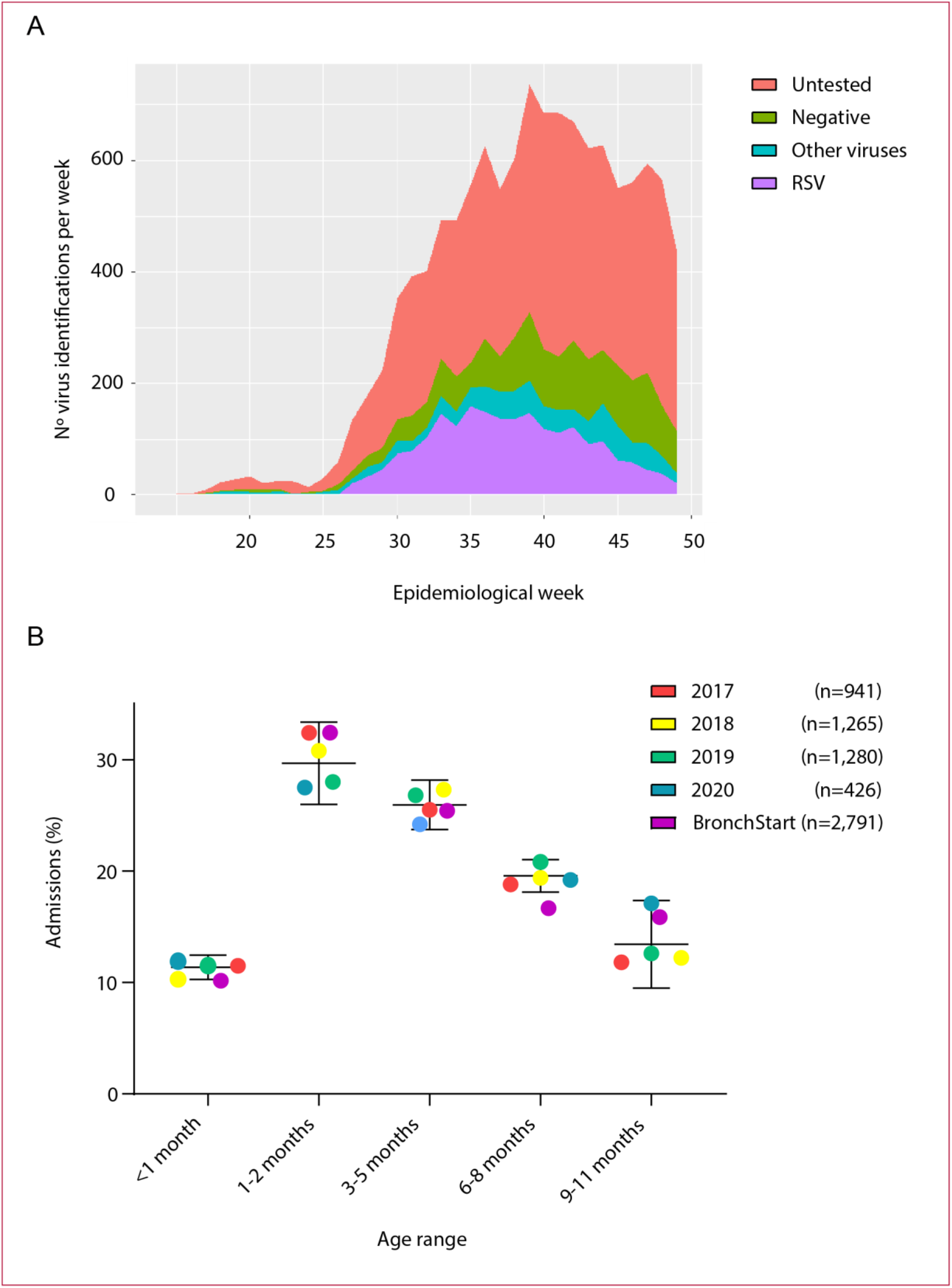
2021 RSV Season in the United Kingdom and Ireland. Virus positivity for BronchStart participants (A), showing peak in rates of RSV positivity between epidemiological weeks 30 and 40. Age at admission for previous 2017-2020 at Leicester Children’s Hospital and Bristol Royal Hospital for Children compared to BronchStart participants (B). Mean and 95% confidence intervals for 2017-2020 admissions shown for each age category, together with total number of infants for year.

Unlike New Zealand, the overall hospital burden of bronchiolitis in the UK and Ireland in 2021 was lower than previous seasons [4]. Disease severe enough to require intensive care was 2.5% in our cohort (for those aged 6 weeks to one year), comparable to 4.2% reported in the BIDS trial [5] (odds ratio using Fisher’s exact test 0.59, 95% confidence interval 0.31-1.18, p = 0.09). We noted a low probability of a SARS-CoV-2-positive RT-PCR test (83/4,328 children tested,1.9%; of which 39 were co-infections with another virus) in children presenting with acute lower respiratory tract infection.

We also observed a substantial number of PED visits and admissions for RSV-positive 12-23 month old children in BronchStart: 362 out of 1,468 (24.7%) admissions were in this age group. This observation of frequent RSV disease in children whose lack of RSV exposure resulted from the delayed seasonal epidemic, should support future long-term follow-up of children born to mothers who receive RSV vaccination in the future. It is possible that waning immunity after initial protection in the period immediately after birth may expose older children to a more severe first episode of RSV disease.

Here we demonstrate the utility of a large, multi-centre study in rapidly gathering data on novel or disrupted infectious disease transmission dynamics requiring a rapid research response. We found that the 2021 summer lower respiratory tract infection peak in the UK and Ireland, although temporally disrupted and with an attenuated disease burden, predominantly affected younger age groups as in previous years. The overall lower burden of disease in 2021 suggests incomplete infection by RSV of its usual susceptible population, probably due to the effect of ongoing NPIs over the study period, and raises the strong possibility of a further wave of infection in the coming months. We suspect that ourimmunity debt has not yet been fully repaid.

## Contributions

Damian Roland: Conceptualization, Methodology, Project Administration, Writing – Original Draft Preparation, Writing – Review & Editing

Thomas C. Williams: Conceptualization, Methodology, Writing – Original Draft Preparation, Writing – Review & Editing

Mark D. Lyttle: Conceptualization, Methodology, Project Administration, Software, Writing –

Original Draft Preparation, Writing – Review & Editing Robin Marlow: Methodology, Data Management

Pia C. Hardelid: Methodology, Writing – Review and Editing

Ian Sinha: Conceptualization, Methodology, Writing – Original Draft Preparation, Writing – Review & Editing

Olivia V. Swann: Conceptualization, Methodology, Writing – Original Draft Preparation, Writing –

Review & Editing

Abigail Maxwell-Hodkinson: Conceptualization, Methodology, Writing – Review & Editing Steve Cunningham: Conceptualization, Methodology, Project Administration, Writing – Original Draft Preparation, Writing – Review & Editing

## Acknowledgements

We thank Khalil Abudahab, Anthony Underwood, and David Aanensen at Microreact for support increating the dashboard, and Linda Wijlaars for assistance in data extraction. We thank Mai Baquedano for technical support in the launch of the REDCap survey tool and ongoing data management, and Darren Goble for information management and technology support, including maintenance of the server and development of a data flow pipeline for the BronchStart outputs. We thank Elizabeth Whittaker for input at the project planning stage. We thank the RESCEU investigators for their support.

## Data availability

Data from the BronchStart Study has been made openly available on a dashboard created by Microreact (https://tinyurl.com/Bronch-Start).

## Supplementary material

Supplementary File 1 lists the PERUKI investigators who have been recruiting patients to the BronchStart Study.

## Ethics

This study has been registered with the NIHR (Research Ethics Committee number 21/HRA/1844)and clinicaltrials.gov(Identifier NCT04959734).

