## Supplementary File 1 for "Features of an aseasonal 2021 RSV epidemic in the UK and Ireland: analysis of the first 10,000 patients"

PERUKI Site Leads and Co-leads

| Site | Country | City | Site lead | Co-lead |
|---|---|---|---|---|
| Alder Hey Children's Hospital NHS Foundation Trust | England | Liverpool | Meriel Tolhurst-Cleaver | |
| Birmingham Children's Hospital | England | Birmingham | Stuart Hartshorn | |
| Bolton NHS Foundation Trust | England | Bolton | Jessica Watson | |
| Bristol Royal Hospital for Children | England | Bristol | Roisin Begley | |
| Chelsea and Westminster NHS Foundation Trust | England | London | Sakura Hingley | Manali Dutta, Gemma Ramsden |
| Children's Health Ireland at Crumlin | Ireland | Dublin | Eleanor Ryan | |
| Children's Health Ireland at Tallaght | Ireland | Dublin | Sheena Durnin | Stanley Koe |
| Countess of Chester NHS Foundation Trust | England | Chester | Steve Brearey | |
| Croydon University Hospital | England | Croydon | Darren Ranasinghe | |
| East Cheshire NHS Trust | England | Macclesfield | Mudiyur Gopi | |
| Frimley Park Hospital | England | London | Patrick Aldridge | Vicky Owens |
| Hull Royal Infirmary | England | Hull | Simon Richardson | |
| Ipswich Hospital | England | Ipswich | David Hartin | |
| John Radcliffe Hospital | England | Oxford | Jiske Steensma | Sahana Rao |
| Leicester Royal Infirmary | England | Leicester | Damian Roland | |
| Leighton Hospital | England | Crewe | Jo Tillett | Simon Dowson |
| Medway Hospital NHS Foundation Trust | England | Gillingham | Adebayo Da Costa | Alfred Sime |
| Newham University Hospital | England | Newham | Claire Kirby | |
| North Middlesex Hospital | England | London | Adam Lawton | |
| Nottingham University Hospitals NHS Trust | England | Nottingham | Ruth Wear | Christopher Gough |
| Ormskirk & District General Hospital | England | Ormskirk | Sharryn Gardner | Craig Rimmer |
| Poole Hospital | England | Poole | Heather Deall | |
| Queen Elizabeth Hospital, Woolwich | England | London | Sharon Hall | |
| Royal Aberdeen Children's Hospital | Scotland | Aberdeen | Catriona Middleton | |
| Royal Alexandra Children's Hospital | England | Brighton | Emily Walton | Friyana Dastur Mackenzie |

| | | | | |
|---|---|---|---|---|
| Royal Berkshire NHS Foundation Trust | England | Reading | Manish Thakker | |
| Royal Derby Hospital | England | Derby | Gisela Robinson | Graham Johnson |
| Royal Hospital for Children, Glasgow | Scotland | Glasgow | Steve Foster | |
| Royal Hospital for Children & Young People, Edinburgh | Scotland | Edinburgh | Jen Browning | Lynsey Rooney |
| Royal Wolverhampton NHS Trust | England | Wolverhampton | Lorna Bagshaw | |
| Salisbury NHS Foundation Trust | England | Salisbury | Seb Gray | |
| Sheffield Children's NHS Foundation Trust | England | Sheffield | Sally Gibbs | |
| South Tyneside & Sunderland NHS Foundation Trust | England | Sunderland | Niall Mullen | |
| Southampton Children's Hospital | England | Southampton | Jane Bayreuther | |
| St George's Hospital, London | England | London | Heather Jarman | |
| St Helens & Knowsley NHS Trust | England | Rainhill | Clare O'Leary | |
| The Royal London | England | London | Raine Astin-Chamberlain | |
| University Hospital Crosshouse | Scotland | Kilmarnock | Lawrence Armstrong | Joanne Mulligan |
| University Hospital Lewisham | England | London | Sophie Keers | |
| Watford General Hospital (West Herts NHS Trust) | England | Watford | Richard Burridge | |
| Wexham Park Hospital | England | Slough | Sarah Wilson | |
| Whipps Cross Hospital | England | London | Amutha Anpananthar | |
| Wirral University NHSFT | England | Birkenhead | David Lacy | |
